# A Bayesian nonparametric method for detecting rapid changes in disease transmission

**DOI:** 10.1101/2022.07.04.22277234

**Authors:** Richard Creswell, Martin Robinson, David Gavaghan, Kris V Parag, Chon Lok Lei, Ben Lambert

## Abstract

Whether an outbreak of infectious disease is likely to grow or dissipate is determined through the time-varying reproduction number, *R*_*t*_. Real-time or retrospective identification of changes in *R*_*t*_ following the imposition or relaxation of interventions can thus contribute important evidence about disease transmission dynamics which can inform policymaking. Here, we present a method for estimating shifts in *R*_*t*_ within a renewal model framework. Our method, which we call EpiCluster, is a Bayesian nonparametric model based on the Pitman-Yor process. We assume that *R*_*t*_ is piecewise-constant, and the incidence data and priors determine when or whether *R*_*t*_ should change and how many times it should do so throughout the series. We also introduce a prior which induces sparsity over the number of changepoints. Being Bayesian, our approach yields a measure of uncertainty in *R*_*t*_ and its changepoints. EpiCluster is fast, straightforward to use, and we demonstrate that it provides automated detection of rapid changes in transmission, either in real-time or retrospectively, for synthetic data series where the *R*_*t*_ profile is known. We illustrate the practical utility of our method by fitting it to case data of outbreaks of COVID-19 in Australia and Hong Kong, where it finds changepoints coinciding with the imposition of non-pharmaceutical interventions. Bayesian nonparametric methods, such as ours, allow the volume and complexity of the data to dictate the number of parameters required to approximate the process and should find wide application in epidemiology.

**Highlights:**

- Identifying periods of rapid change in transmission is important for devising strategies to control epidemics.
- We assume that the time-varying reproduction number, *R*_*t*_, is piecewise-constant and transmission is determined by a Poisson renewal model.
- We develop a Bayesian nonparametric method, called EpiCluster, which uses a Pitman Yor process to infer changepoints in *R*_*t*_.
- Using simulated incidence series, we demonstrate that our method is adept at inferring changepoints.
- Using real COVID-19 incidence series, we infer abrupt changes in transmission at times coinciding with the imposition of non-pharmaceutical interventions.

## 1. Introduction

Throughout the SARS-CoV-2 pandemic, the *time-varying* reproduction number^1^, *R*_*t*_, has been estimated and used to gauge the effectiveness of control measures (e.g. Flaxman et al. (2020); Li et al. (2021); Parag et al. (2021); Brauner et al. (2021); meta-analysis of such studies: Mendez-Brito et al. (2021)). *R*_*t*_ represents the average number of secondary cases spawned by a single primary case. When *R*_*t*_ *>* 1, an outbreak is expected to grow exponentially; public health interventions try to permanently shift *R*_*t*_ *<* 1 meaning an epidemic will, in the long run, die out.

A widely used approach for estimating *R*_*t*_ is through *renewal equations* which assume that future numbers of cases depend on the history of case counts, the generation times, representing the typical timescales between primary and secondary infections, and *R*_*t*_ (theory: Fraser (2007); Nishiura and Chowell (2009); example software: Thompson et al. (2019)). These models are typically formulated in discrete time (usually at the daily resolution), and the dynamics are assumed stochastic. Here, we focus on the most popular version of these models which assume that the population is well-mixed and that there is no demographic heterogeneity.

A variety of approaches exist for estimating *R*_*t*_ using time series incidence data, either in real-time (i.e. using only information up until a current time *t*; Cori et al., 2013; Parag, 2021) or retrospectively (Wallinga and Teunis, 2004). These approaches make diverse assumptions about the continuous structure of *R*_*t*_: that it is piecewise-constant within a sliding window of a given prespecified length (Wallinga and Teunis, 2004; Thompson et al., 2019); that it varies smoothly with the variation controlled by a Gaussian filter (Abbott et al., 2020; Parag, 2021); or that it is made up of an inferred number of pieces with a single, optimal, number of pieces inferred by considering a criterion derived from information theory (Parag and Donnelly, 2020).

Our approach also assumes that *R*_*t*_ is piecewise-constant, and that, within each piece, the epidemic follows a standard Poisson renewal process (Cori et al., 2013). We do not specify the number of pieces nor provide a limit on this number *a priori*. To do so, we use a Bayesian model with a *Pitman-Yor process* prior (Pitman and Yor, 1997) to represent the values of *R*_*t*_ across any feasible number of pieces. This process comes from the field of Bayesian nonparametrics—a broad class of models where the data are modelled by a (potentially) countably infinite set of parameters, where the complexity of the models, indexed by the number of parameters, increases in lockstep with the volume and complexity of the data (Ghahramani, 2013). Our approach, which we call *EpiCluster*, avoids the need to directly specify how often and how fast *R*_*t*_ need change to represent a given incidence curve. Instead, the data and a prior jointly determine how many pieces are needed to approximate the *R*_*t*_ curve, and, in §2, we introduce a default prior meant to find a parsimonious approximation of it with few changepoints. Our method, being Bayesian, provides a measure of uncertainty in both the number of pieces and *R*_*t*_ (see Figure 1). We develop an efficient Markov chain Monte Carlo (MCMC) inference method for fitting our model to incidence data using collapsed Gibbs sampling (Lambert, 2018, Chapter 14), which efficiently steps between models of different dimensionalities (corresponding to different numbers of *R*_*t*_ pieces). We provide an open-source Python package implementing EpiCluster, which computes *R*_*t*_ profiles and runs in seconds to minutes (dependent on the length of data series and complexity of the *R*_*t*_ profile), which is available at github.com/SABS-R3-Epidemiology/epicluster.

**Figure 1:**
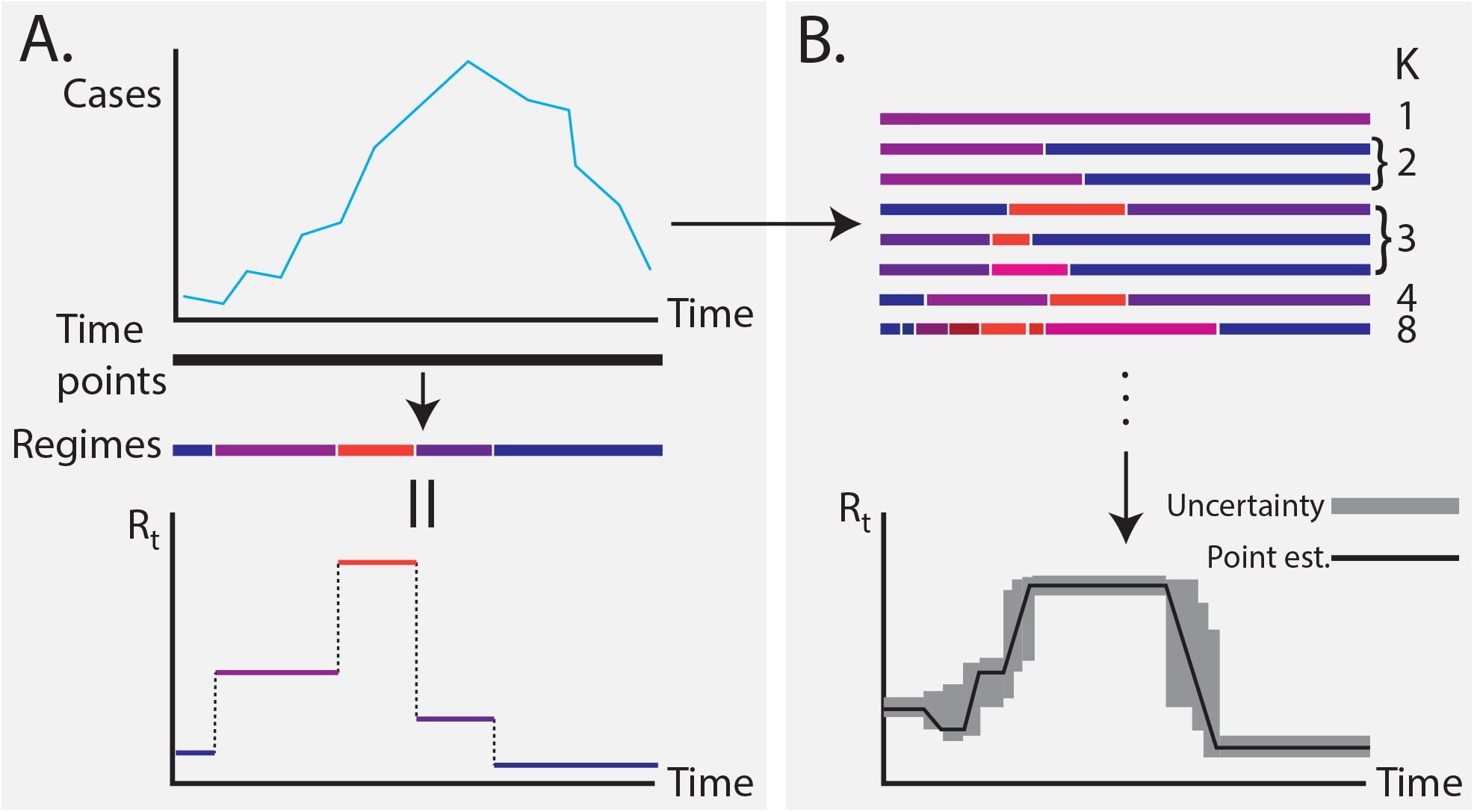
Pitman-Yor based inference for the time-varying reproduction number. Panel A represents the intrinsic assumption underpinning our method: that R_*t*_ is piecewise-constant, and the pieces are shown as different coloured bars. Panel B shows how our nonparametric prior allows a decomposition of the time points into partitions comprising different numbers of pieces (K). Our MCMC sampler (see Algorithm 1) explores this space over partitions efficiently, resulting in posterior uncertainty in R_*t*_.

By fitting our model to simulated data with known *R*_*t*_ profiles (in §3.1), we show that EpiCluster is adept at identifying times of rapid change in *R*_*t*_ as may occur following the imposition of major and broad-scale interventions (Dehning et al., 2020; Flaxman et al., 2020; Brauner et al., 2021)—either in real-time or retrospectively. It is less well suited to estimate *R*_*t*_ if it changes more gradually, and more appropriate methods exist for this purpose (e.g. Thompson et al., 2019; Parag, 2021). Unlike methods which directly model *R*_*t*_ as a function of known intervention timings and severities (e.g. Dehning et al., 2020; Flaxman et al., 2020; Brauner et al., 2021), our method is purely driven by the incidence series. Because of this, it provides a straightforward and intervention-agnostic initial step for assessing the impact of interventions, and similarly agnostic approaches have previously been used in retrospective analyses of COVID-19 transmission (Parag et al., 2021). Since it does not use additional information about interventions, our approach is likely to produce estimates with greater variability. But, it requires fewer assumptions to be made, which may be beneficial, since the assumptions around intervention timing (Soltesz et al., 2020) and modelling details (Sharma et al., 2020) may affect estimates and their interpretation. In §3.4, we apply our framework to data from the COVID-19 outbreaks in Australia and Hong Kong and show that it is able to find changepoints in *R*_*t*_ corresponding to the imposition of known interventions. Our method provides a tool for outbreak analysis complementary to existing methods and could form part of an analysis pipeline for associating interventions with changes in transmission.

## 2. Methods

### 2.1. Renewal process model

We estimate *instantaneous reproduction numbers* and mean this whenever we write *R*_*t*_. Instantaneous reproduction numbers represent the average number of secondary cases that would be generated by an infected case at time *t* assuming that future transmission remains the same as at time *t* (Fraser, 2007). We assume that the data consist of a series of daily case counts^2^ for each day, *t*, from *t* = 1 to 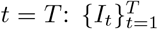 and that the case counts are perfectly known. Due to the within- and and between-individual variability in rates of contact and infectivity, a *generation time distribution* is used to represent the duration between the time at which a parent case occurs and its offspring. We model the case count *I*_*t*_ as arising according to the Poisson renewal process:

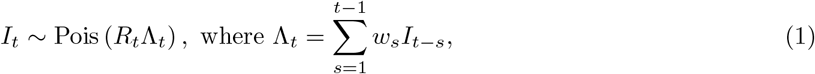

where *R*_*t*_ ≥ 0 is the time-varying reproduction number on day *t*, and Λ_*t*_ ≥ 0 is the transmission potential. The *w*_*s*_ terms represent the generation time distribution: 0 ≤ *ω*_*s*_ ≤ 1 indicates the probability that a primary case takes between *s* − 1 and *s* days to generate a secondary case, and 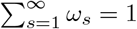. Since it is not typically possible to know when individuals become infectious, generation times are not directly observed, making it difficult to estimate the generation time distribution. Here, we use the *serial interval distribution* in its place, which describes the time between the onset of symptoms between a primary and secondary cases. This is easier to estimate from infector-infectee pairs since it is more directly observable and has a similar mean (Svensson, 2007).

### 2.2. Model of changing R_t_

#### 2.2.1. Exchangeable partition probability functions and the Pitman-Yor process

Here, we assume that the *R*_*t*_ profile can be decomposed into a number of regimes within which *R*_*t*_ is constant. Our goal is to avoid prespecifying the location of changepoints—representing the boundary between two different *R*_*t*_ regimes—nor their count, since these choices can bias analyses, but rather to learn an appropriate configuration of the time points into regimes using Bayesian inference. We develop a probabilistic model of the division of the time points into regimes. To do so, we use a Pitman-Yor process (Pitman and Yor, 1997)^3^ to account for a probabilistic decomposition of data points into clusters and, following Martínez and Mena (2014), we adjust this model to account for the time series nature of our data. The remainder of this subsection serves as a brief review of this model, starting with a treatment of the nonparametric clustering of unordered data points via *exchangeable partition probability functions* (EPPFs) and followed by appropriate modifications for the time series case (see §2.2.2).

In the standard clustering problem, we have a set [*T*] = {1, …, *T* } (i.e., the labels of *T* data points), which we would like to divide into *K* mutually exclusive subsets {*A*_1_, …, *A*_*K*_} such that U_*k*_*A*_*k*_ = [*T*] where none of the *A*_*k*_ are empty. We denote the set of all such groupings by 𝒫_[*T*]_; each element of 𝒫_[*T*]_ is called a *partition*. Random variables Π_*T*_ taking values in 𝒫_[*T*]_ are termed *random partitions* of [*T*]. A random partition has the property of *exchangeability* if its probability distribution can be written as a symmetric function *p* of the subset sizes, i.e.,

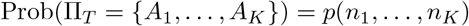

where *n*_*k*_ = |*A*_*k*_| (i.e. *n*_*k*_ is the size of the subset, *A*_*k*_).

Under these conditions *p* is known as an EPPF. A more complete treatment of the concept of EPPFs can be found in Pitman (2002); Lijoi and Prunster (2010). A fairly general EPPF, which we will employ in this work, is derived from the Pitman-Yor process, a generalisation of the Dirichlet process (Teh, 2010). This EPPF is given by (Pitman, 2002, eq. (3.6)):

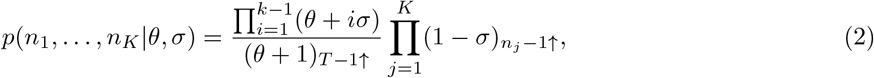

where 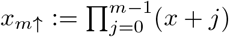, and *σ* ∈ [0, 1) and *θ >* − *σ* are the two hyperparameters governing the process: *σ* is called the discount parameter, which essentially controls how the number of regimes, *K*, grows with the size of the dataset; *θ* is called the strength parameter with larger values giving greater weight to series with more regimes. In the limit *σ* → 0, a Pitman-Yor process becomes a Dirichlet process which permits a slower growth (of order log *T* opposed to *T* ^*σ*^; Pitman, 2002, section 3.3) in the number of regimes with increases in data size.

#### 2.2.2. Applicability of EPPFs to time series problems

Unlike the general clustering problem, in the time series case, the data points have an ordering which the clusters must respect. For example, consider an incidence series of length three: (*I*_1_, *I*_2_, *I*_3_). For this series, allowable effective reproduction number allocations include: {{*I*_1_, *I*_2_, *I*_3_}}, where all the data points are generated from a process with the same effective reproduction number: i.e. there is a single regime (*K* = 1); {{*I*_1_}, {*I*_2_, *I*_3_}}, where the first data point was generated from a process with one effective reproduction number and the latter two data points from a process with a different one: i.e. there are two regimes (*K* = 2); {{*I*_1_, *I*_2_}, {*I*_3_}}, where the first two points are grouped; and {{*I*_1_}, {*I*_2_}, {*I*_3_}}, where each data point is generated from a process with a different reproduction number: i.e. there are three regimes (*K* = 3).

An allocation which would be disallowed is: {{*I*_1_, *I*_3_}, {*I*_2_}}, where the first and third data points come from the same process which is distinct from that governing the second. Whilst, it is possible that transmission could return to a previous level, it is an assumption of our modelling process that only consecutive data points share the same *R*_*t*_. By avoiding recurrence to historical regimes, we ensure that the changepoints identified are straightforward to interpret.

For a given EPPF, *p*^′^, we can obtain a distribution *p* which is supported only on those partitions which respect an ordering of the labels using the following result (Martínez and Mena, 2014):

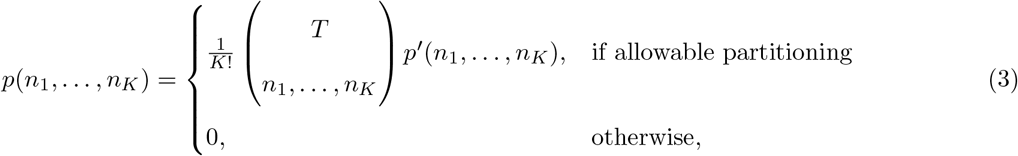

where the large bracketed term indicates the multinomial coefficient.

Combining eqs. (3) and (2), we obtain the following result for the prior distribution on the sequence of regime sizes in the time series case:

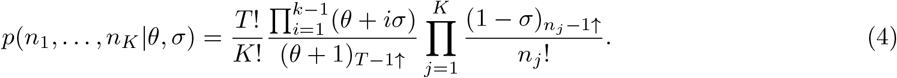

#### 2.2.3. Hyperparameters of the process

In order to learn parsimonious assignments of the time points into regimes, our prior, given by eq. (4), should favour configurations consisting of longer regimes. We favour longer regimes because they mitigate against overfitting—for typical data, the likelihood of the renewal process would be maximised by assigning each time point to its own cluster with an idiosyncratic value of *R*_*t*_; the resulting profile of *R*_*t*_ values will tend to be jagged and exhibit spurious fluctuations. Additionally, longer regimes have the advantage of allowing more data to be leveraged in order to learn more precise estimates of *R*_*t*_. However, by favouring longer regimes, it is possible that we miss shorter term fluctuations in *R*_*t*_—this is akin to the issue of choosing window lengths for a number of existing methods (e.g. Thompson et al., 2019).

Eq. (4) induces a marginal distribution over the number of clusters whose mean has been derived as (Pitman, 2002, eq. (3.13)):

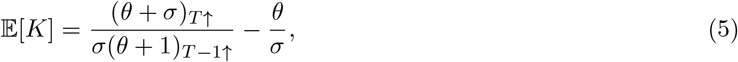

for *σ* ≠ 0. For small values of the hyperparameters *θ* and *σ*, 𝔼 [*K*] is significantly smaller than the number of time points *T* (see Fig. S1), and the marginal distribution of *K* places little weight on values of *K* close to *T*, thus preferring sparsity in the number of clusters. For all results presented in this paper, we set *θ* = 0 and choose *σ* as a function of *T* such that *E*[*K*] = 1.5 (with the appropriate value of *σ* selected by numerical optimization of eq. (5)); this represents a prior belief that *R*_*t*_ is generally constant over the time series, but allows flexibility to add clusters when the data provides evidence that they are needed. For a time series of length *T* = 100, our choice of prior hyperparameters induces a marginal distribution over the number of clusters whose 2.5^th^ percentile is 1 cluster and 97.5^th^ percentile is 4 clusters.

### 2.3. Marginal likelihood of the data

In this subsection, we calculate the marginal likelihood of the data conditional on a particular arrangement of the time points into regimes, which involves integrating out *R*_*t*_ with respect to its prior distribution. This marginal likelihood enables efficient inference for the posterior distribution over regime configurations via collapsed Gibbs sampling (see § 2.4).

The marginal likelihood for an incidence series conditional on a particular set of subset sizes *n*_1_, …, *n*_*K*_ (see §2.2) can be written as a product of marginal likelihoods for each regime:

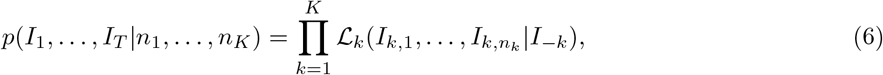

where *I*_*k,j*_ denotes the *j*th data point in regime *k*, and ℒ_*k*_ is the marginal likelihood of the data in the *k*th regime, which we assume is conditional on all cases observed prior to regime *k* (denoted by *I*_−*k*_). We derive the regime-specific marginal likelihoods using the renewal model (eq. (1)):

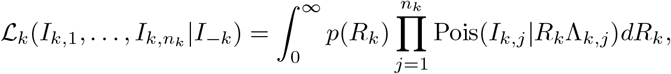

where Λ_*k,j*_ is the transmission potential calculated for the *j*th time point in regime *k, R*_*k*_ is the value of the effective reproduction number for the *k*th regime, and *p*(*R*_*k*_) is the prior on *R*_*k*_.

We choose a gamma distribution prior for *R*_*k*_ with shape parameter *α* and rate parameter *β*.^4^ With this choice of prior, the integral in the formula for the regime-specific marginal likelihood can be evaluated analytically, resulting in:

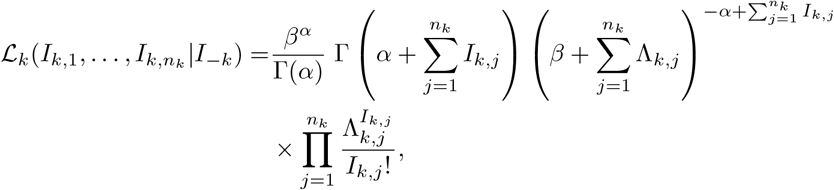

where Γ(·) is the gamma function.

Additionally, with the gamma prior on *R*_*k*_, the posterior distribution of each *R*_*k*_, conditional on the data assigned to regime *k*, is given by the conjugate gamma posterior (Creswell et al., 2022):

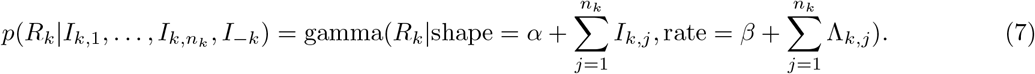

As prior hyperparameters, we select *α* = 1 and *β* = 0.2. With this choice, the prior mean and standard deviation are both equal to 5. The high standard deviation provides a relatively uninformative prior, and the high mean ensures that the outbreak is unlikely to be determined as under control (since *>* 81% of prior probability is for *R*_*t*_ *>* 1) unless there is considerable evidence to suggest otherwise.

### 2.4. Inference

At particular values of the hyperparameters *σ* and *θ*, the target posterior of regime configurations, which we denote by *p*(*n*_1_, …, *n*_*K*_|*I*_1_, …, *I*_*T*_, *σ, θ*) is proportional to the product of and eq. (4) and eq. (6):

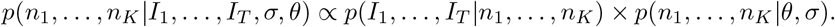

For brevity, we suppress the dependence on cases and hyperparameters and denote the unnormalized posterior by *p*(*γ*_*K*_), where *γ*_*K*_ := (*n*_1_, …, *n*_*K*_) indicates a particular configuration of the time points into *K* regimes.

Inference for this posterior is performed via Markov Chain Monte Carlo (MCMC) which provides a distribution over the number of regimes by jumping between models of different numbers of parameters. We use the same split-merge-shuffle structure as Martínez and Mena (2014). Each step of our MCMC algorithm is given in Algorithm 1, and we now describe it.

Different configurations of the time points into regimes are explored through the use of *split, merge*, and *shuffle* proposals. The split proposal takes an existing regime and proposes to split it into two regimes at some randomly located changepoint. The merge proposal takes two consecutive regimes and proposes to merge them into one. Both of these proposals consider an update to the total number of regimes, thus allowing the sampler to explore the marginal posterior distribution over the number of regimes. Additionally, the shuffle proposal shifts the boundary between two consecutive regimes, thus keeping the same number of regimes but efficiently exploring uncertainty in the location of a changepoint. At each iteration of the MCMC sampler, we make one shuffle proposal and randomly choose whether to make a split or merge proposal, with the MCMC tuning parameter *q* giving the probability of making the split proposal. For the results presented in this paper, we fix *q* = 0.5. The acceptance probabilities for the split, merge, and shuffle proposals are derived in Martínez and Mena (2014) and are given by min(1, *α*_*e*_), with *e* ∈ {split, merge, shuffle}.

*α*_split_ is calculated by:

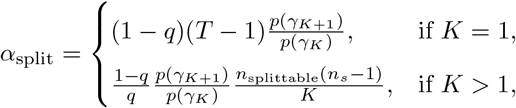

where *n*_splittable_ is the number of splittable regimes (i.e., those with more than one time point assigned to them) in the original configuration, and *n*_*s*_ is the length of the regime selected for a split; *γ*_*K*_ is the current regime configuration, and *γ*_*K*+1_ is the split configuration.

The corresponding quantity for a merge move is given by:

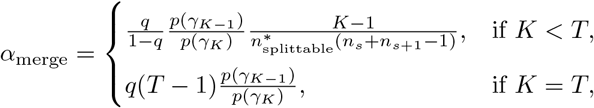

where 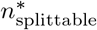 is the number of splittable regimes in the proposed configuration, and *n*_*s*_ and *n*_*s*+1_ are the sizes of the regimes which are proposed to be merged; *γ*_*K*−1_ is the merged regime configuration.

The equivalent quantity for a shuffle move is given by:

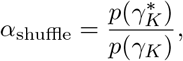

where 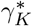 is the shuffled configuration obtained from *γ*_*K*_ as described in Algorithm 1.

The values of *R*_*t*_ are updated using Gibbs steps conditional on the current regime configuration. We run four separate MCMC chains, two initialised with all time points assigned to a single regime (i.e. *K* = 1) and the other two initialised with all time points assigned to their own singleton regime (i.e. with *K* = *T*). We assessed convergence of our MCMC algorithm (Algorithm 1) by monitoring convergence in *K*, the number of regimes. To do so, we computed the 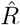 statistic (Gelman and Rubin, 1992) and required 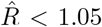. Once convergence was determined, we discarded the first 50% of each of the MCMC chains as warm-up and combined the rest of the samples in order to calculate posterior percentiles and means.

#### Algorithm 1 One step of the MCMC sampler.

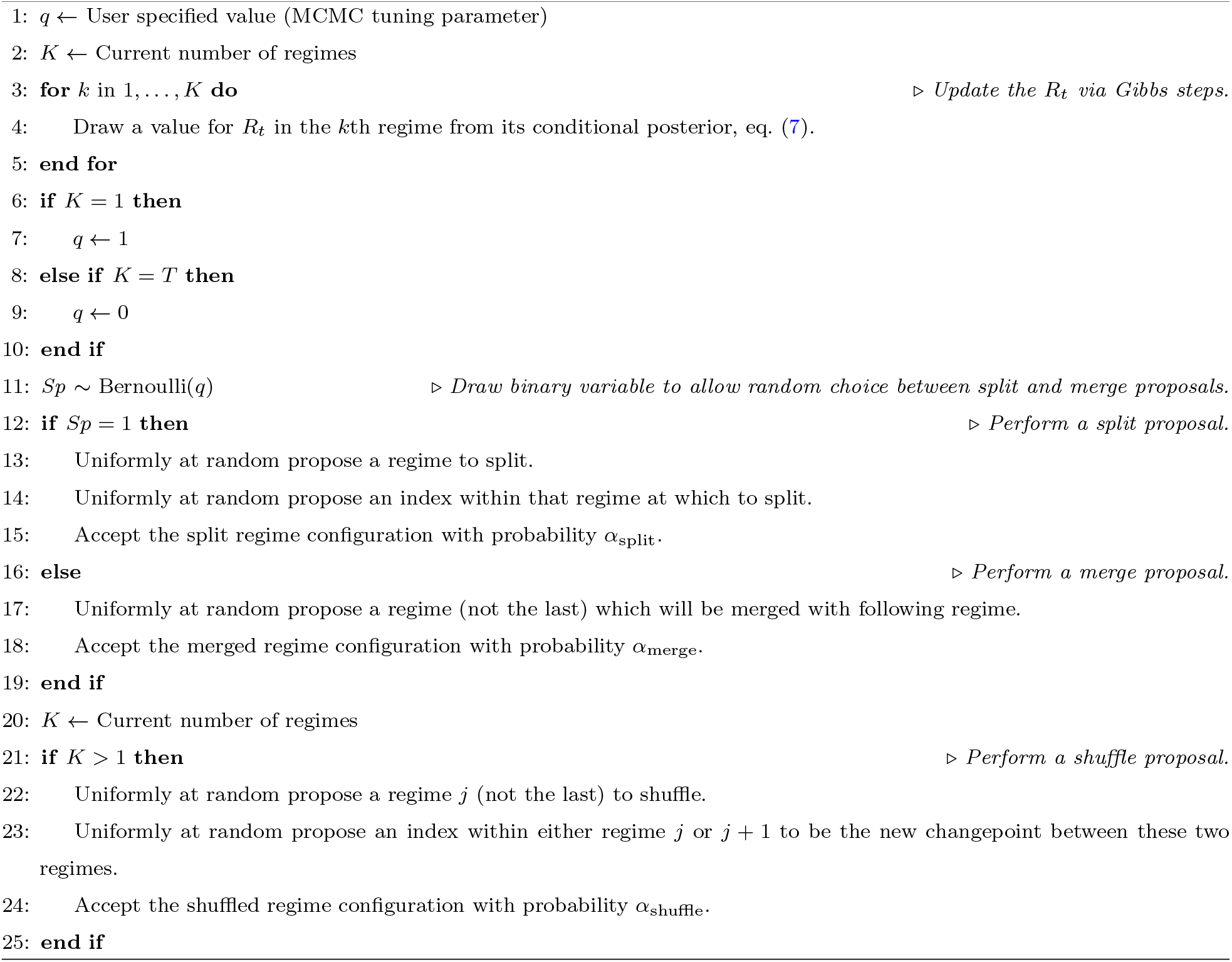

### 2.5. Comparator methods

In § 3, we compare the posterior distribution for *R*_*t*_ yielded by our nonparametric method to those yielded by two comparator methods. This first is the Cori sliding window method (Cori et al., 2013; Thompson et al., 2019), which assumes that *R*_*t*_ is constant over a sliding window of *τ* days looking backwards. The sliding window width has a significant effect on the posterior and the effective bias-variance trade-off. As a result, we consider two choices of *τ* (7 days and 28 days) when applying the method to synthetic data. The second comparator is the EpiFilter method (Parag, 2021), which applies sequential Bayesian smoothing and controls change in *R*_*t*_ under a random walk prior.

### 2.6. Implementation and runtime

We implemented EpiCluster in Python 3. A Python package of the model, including the MCMC inference algorithm, is available at github.com/SABS-R3-Epidemiology/epicluster, while the notebooks and data for reproducing all results in this paper are available at github.com/SABS-R3-Epidemiology/epicluster-results. We ran the sliding window method using the **branchpro** Python package (Creswell et al., 2022). We ran the EpiFilter method through its R package (Parag, 2021). Using our software library and typical consumer hardware (3.6GHz CPU), EpiCluster takes from several seconds to several minutes to learn the posterior, depending on the complexity of the *R*_*t*_ profile. By comparison, the sliding window method and EpiFilter methods are effectively instantaneous to compute on the time series studied here.

### 2.7. Handling imported cases

Some of the real data examples we consider (see §3.4) consist of case counts in locations where a substantial proportion of the case loads are due to imported cases. To account for this, we adapt our renewal model using the methods described in Creswell et al. (2022). In this approach, cases are classified as either *local* or *imported*. Local cases 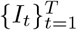 are those arising from local transmission in the spatial region under consideration, while imported cases 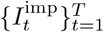 are those who were infected elsewhere before travelling to the region. Thus, imported cases contribute to local transmission, but did not arise from it. In outbreaks where a significant proportion of cases are imported, distinguishing local from imported cases is important for accurate estimation of *R*_*t*_ (Roberts and Nishiura, 2011; Thompson et al., 2019). We allow local and imported cases to have different risks of onwards transmission by weighting the imported cases by some number *ϵ >* 0 (Creswell et al., 2022), and we set *ϵ* to appropriate values (see §2.8). The default choice of *ϵ* = 1 corresponds to an equal risk of onwards transmission between local and imported cases. Note, any case and any subsequent lineages begot by an imported case are classified as local: it is only the rate at which newly imported cases infect others which is assumed to differ from purely local transmission.

We adapt eq. (1) to model the dynamics of local cases *I*_*t*_, resulting in:

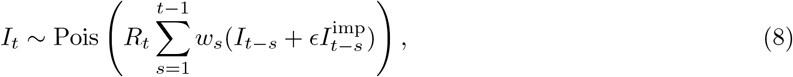

where *R*_*t*_ is the effective reproduction number that characterises local transmission on day *t*. For problems where imported cases are not considered, we use eq. (1).

### 2.8. Real incidence data

We fit to real case incidence data for local and imported COVID-19 cases for three regions: Victoria and Queensland in Australia and Hong Kong. In each of these three locations, we used cases with dates given by the date of symptom onset. We selected these regions as they exhibit a variety of different trends in *R*_*t*_: a gradual decrease in Victoria, a more rapid decrease in Queensland, and a fall in *R*_*t*_ followed by the sudden appearance of a second wave in Hong Kong. Data for the Australian regions were obtained from the Australian national COVID-19 database (Price et al., 2020); data for Hong Kong were obtained from the Hong Kong Department of Health COVID-19 database (Hong Kong Department of Health, 2022). For the Australian states, cases of unknown origin were assumed to be local, and in Hong Kong, all cases other than those listed as “imported case confirmed” were treated as local. We assumed *ϵ* = 1 in eq. (8) for Victora and Queensland; however, for Hong Kong, transmission networks suggest that imported cases were significantly less infective than local cases, so we set *ϵ* = 0.2 (Liu et al., 2021; Creswell et al., 2022). In all three instances, we assumed that under-reporting and delays were negligible given the strong surveillance in these countries.

## 3. Results

### 3.1. EpiCluster reliably estimates sudden changes in R_t_ in retrospective analyses

To evaluate the performance of our model, we generated synthetic incidence data using eq. (1) where the *R*_*t*_ profile was known (see Figure 2). We considered three *R*_*t*_ profiles: one with a precipitous decline in *R*_*t*_ (“fast drop-off”); another, with a decline in *R*_*t*_ followed by a later resurgence (“fast resurgence”; we included this profile since resurgences are harder to infer than declines in transmission strength; Parag and Donnelly, 2022); and another with a more gradual decline in *R*_*t*_ (“slow drop-off”). The fast drop-off and slow drop-off time series were initialized with 5 cases on each of three days preceding the beginning of simulation, while the fast resurgence was initialized with 5 cases on each of fifty days preceding the beginning of simulation. Simulations for fast drop-off and slow drop-off used the COVID-19 serial interval (Nishiura et al., 2020), while the fast resurgence used the Ebola serial interval as estimated for the 2014 West African Outbreak (Van Kerkhove et al., 2015).

**Figure 2:**
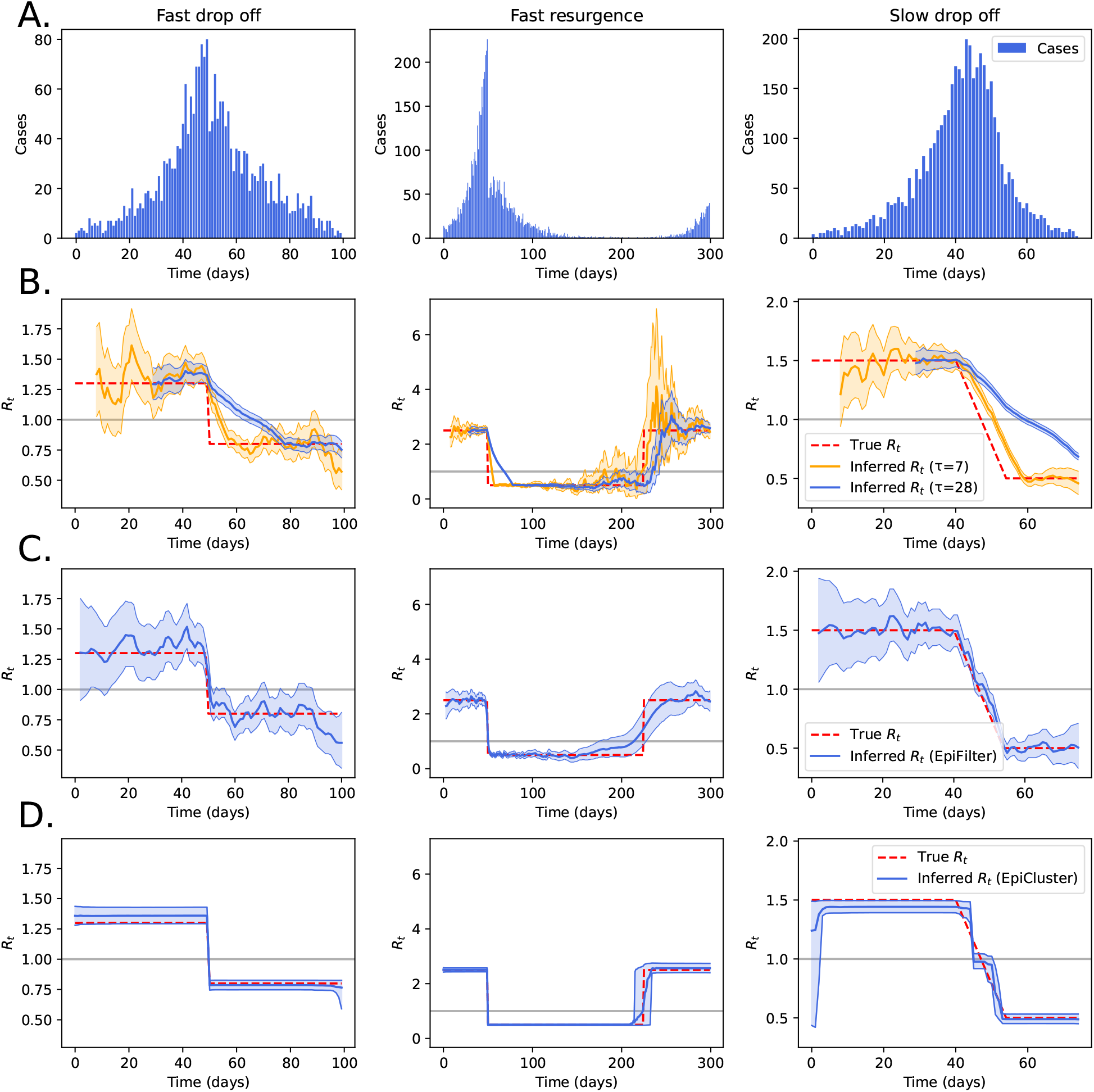
Recovering synthetic R_*t*_ profiles in retrospective analyses. We generated synthetic case data (panel A) using the Poisson renewal model (eq. (1)) with three prespecified profiles for R_*t*_ (dashed red lines in panels B / C / D). In panel B, we show the inferred R_*t*_ profile using a sliding window method (Thompson et al., 2019) for two different choices of the sliding window size (τ = 7 and 28 days). In panel C, we show the inferred R_*t*_ profile using the EpiFilter method (Parag, 2021). In panel D, we show the inference results when using EpiCluster to recover R_*t*_. In panels B, C and D, shaded regions indicate the central 90% of the posterior distribution of R_*t*_, while the central line indicates the posterior mean, and the background gray line indicates R_*t*_ = 1.

In Figure 2, we compare *R*_*t*_ estimates from our method with those from two comparator methods: the sliding window method (Thompson et al., 2019) with two different choices of the sliding window width (7 days and 28 days), and the EpiFilter method (Parag, 2021).

Across the three *R*_*t*_ profiles considered, the estimates from the sliding window method lag behind the true values (Fig. 2B), since the windows are inherently backward-looking—the longer the window width, the longer the moving average and the slower it is to respond to changes in *R*_*t*_; the estimates are also very variable. The EpiFilter method fares better and is able to reliably infer downward shifts in *R*_*t*_ (Fig. 2C), corresponding to suppression; this method overly smooths over the upward tick in transmission in the fast-resurgence example. Our method performs favourably in the two “fast” examples (Fig. 2D). Like the EpiFilter method, our approach is less able to infer resurgence than suppression (Parag and Donnelly, 2022). In the slow example, our piecewise-constant method approximates the linear decline in *R*_*t*_ with a staircase-like profile, which is better estimated by EpiFilter. In Figure S2, we show the effect of changing the hyperparameters of our method on inference for the slow drop-off example on the number and location of the regimes which are learned. As the two hyperparameters, *θ* and *σ* increase, more weight is given to a partitioning consisting of more regimes (see also Fig. 1), and the staircase steps become finer.

To account for stochastic variation in the synthetic data generation, we repeated inference for the fast resurgence example 10 times (Fig. S3). For the three methods, the posterior means are qualitatively similar across all runs, suggesting that these results are consistent across different realisations of the renewal process.

In the fast drop off and fast resurgence examples, EpiCluster estimates *R*_*t*_ with low bias and high precision. This is because the *R*_*t*_ profiles in the simulated examples align well with the assumptions made in our modelling: namely, that the *R*_*t*_ profile is piecewise-constant. We now consider *R*_*t*_ profiles with notable deviations from this assumption. In Figure 3, we compare the same methods on both noisy (left and middle columns) and oscillatory *R*_*t*_ profiles. When the magnitude of the noise is low (left column), the results mirror those from the previous example. When the noise level increases (middle column), all methods are late to predict the precipitous decline in *R*_*t*_, and EpiFilter provides a better quantification of uncertainty than the nonparameteric model. For the sinusoid example (right column), EpiFilter performs best, since the assumptions underpinning that method—that *R*_*t*_ follows a random walk—are closer to the reality of the generated data.

**Figure 3:**
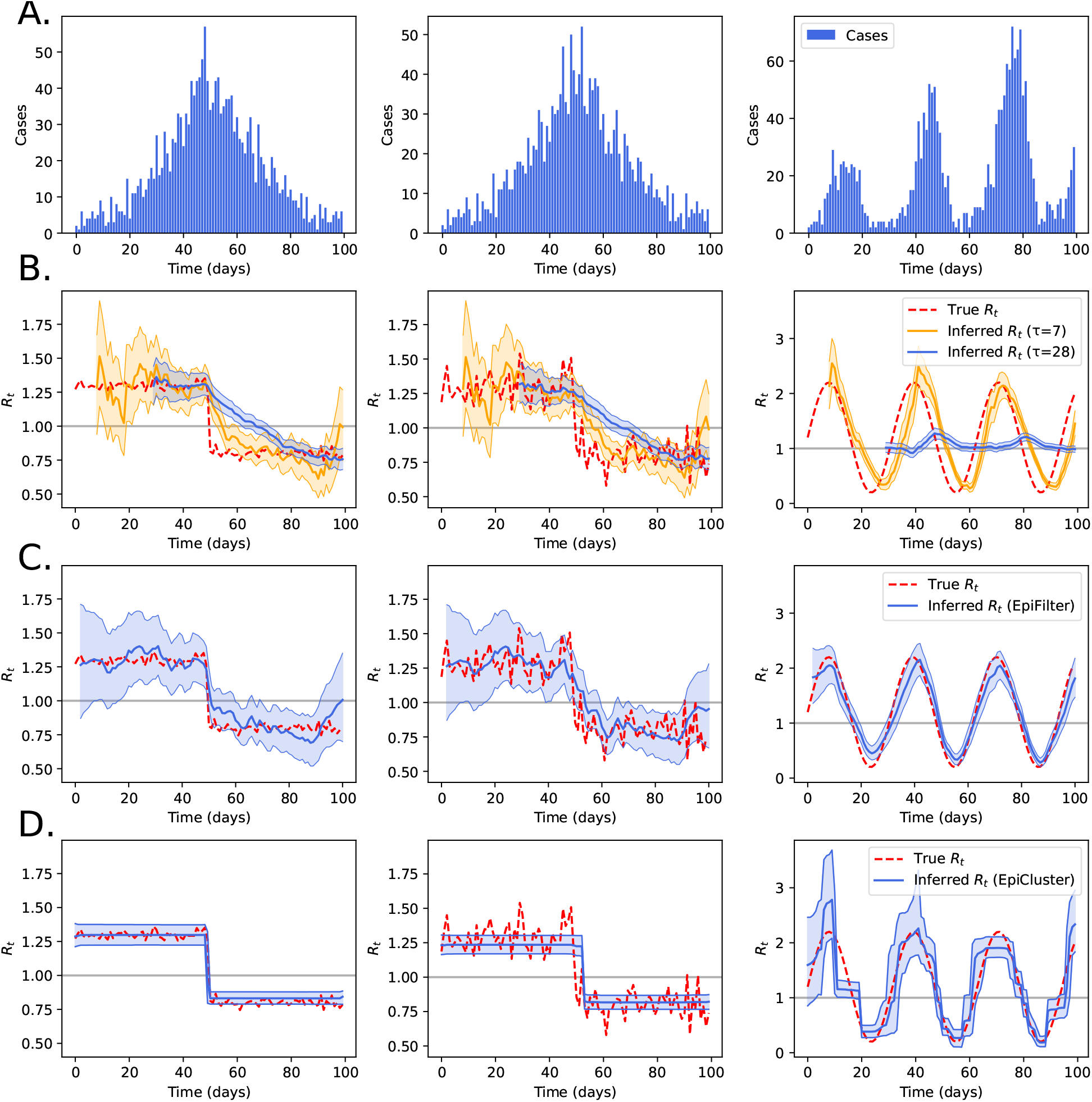
Recovering noisy and oscillatory R_*t*_ profiles in retrospective analyses. We generated synthetic case data (panel A) using the Poisson renewal model with three prespecified profiles for R_*t*_ (dashed red lines in panels B / C / D). The R_*t*_ profiles were calculated using step functions with additive *i.i.d*. Gaussian noise of standard deviation 0.025 (left) and 0.1 (middle). In the right column, we show results when R_*t*_ follows a sine wave. In panel B, we show the inferred R_*t*_ profile using a sliding window method (Thompson et al., 2019) for two different choices of the sliding window size (τ = 7 and 28 days). In panel C, we show the inferred R_*t*_ profile using the EpiFilter method (Parag, 2021). In panel D, we show the inference results when using EpiCluster to recover R_*t*_. In panels B, C and D, shaded regions indicate the central 90% of the posterior distribution of R_*t*_, while the central line indicates the posterior mean, and the background gray line indicates R_*t*_ = 1.

### 3.2. EpiCluster is effective at detecting sharp changes in transmission in real-time

The results thus far have considered retrospective analysis of outbreaks; these analyses are important for understanding the timing and impact of interventions following their imposition (e.g. Flaxman et al. (2020); Brauner et al. (2021)). But, in unfolding epidemics of novels pathogens, it is crucial to know in as close to real time as data allows whether transmission changes rapidly either after an intervention is instituted or after it is discontinued. In this section, we compare how the three *R*_*t*_ estimation methods fared in inferring an epidemic resurgence in real-time: as new case data becomes available subsequent to a jump upwards in transmission. We used the same fast resurgence data as in Fig. 2 and fit each method for a series of datasets of different lengths. Each of these datasets began at the same point (at *t* = 0); the datasets ended at different points. The endpoints ranged from 5 days to 35 days post-resurgence with gaps of 5 days between them.

The posterior means of the inferred *R*_*t*_ series are shown in Fig. 4, while the full posteriors are shown in Fig. S4. The results illustrate that all three methods needed considerable data post resurgence to infer changes in transmission. For each series, EpiCluster generally fared best in inferring the timing and magnitude of resurgence, with the posterior uncertainty interval reliably including the true *R*_*t*_ profile.

**Figure 4:**
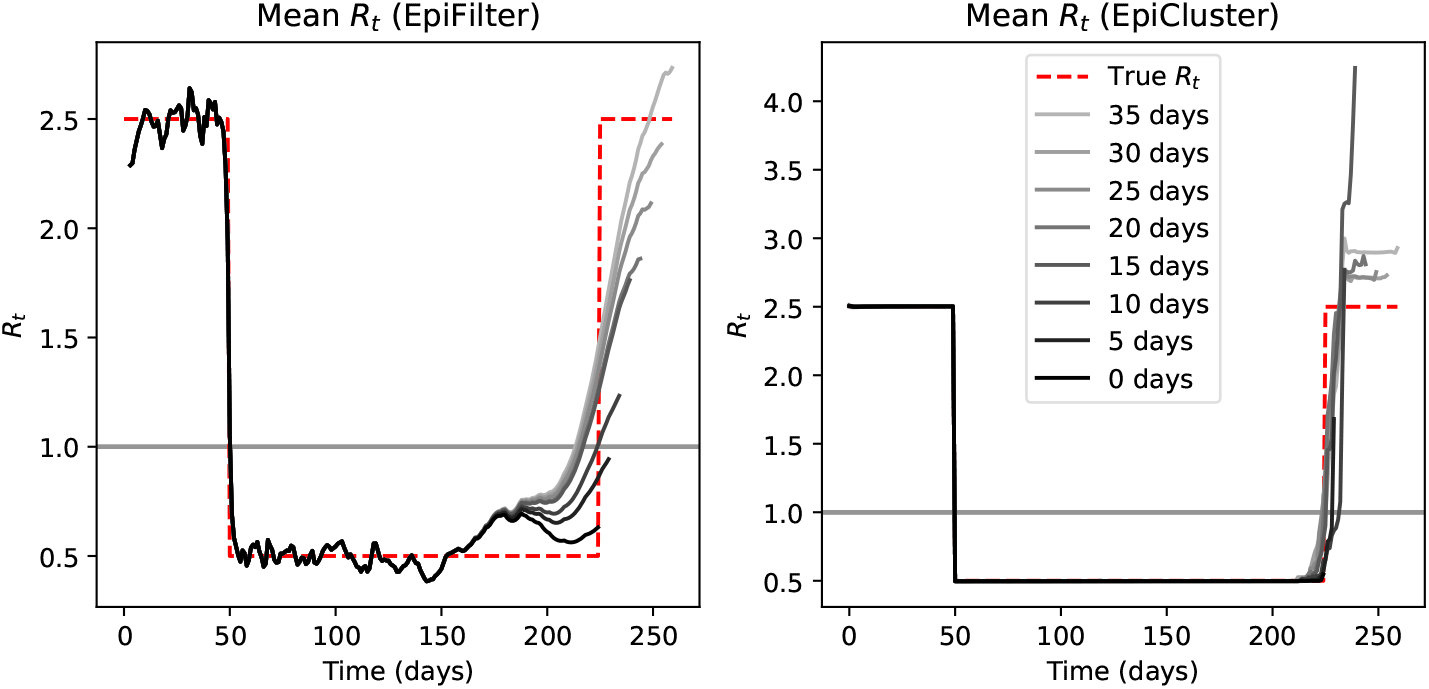
Real-time estimation of a resurgence in R_*t*_. We used the same fast resurgence synthetic data from Figure 2 and performed inference for R_*t*_ based only on the time series up till the number of days after the resurgence indicated in the legend. In the left panel, we show the mean inferred R_*t*_ profile using the EpiFilter method (Parag, 2021). In the right panel, we show the results when using EpiCluster to recover the mean of R_*t*_. The background gray line indicates R_*t*_ = 1.

### 3.3. Data generating processes with greater variability pose issues for all methods and EpiFilter generally performs best

Variation in transmissibility across different individuals within a population can lead to greater variation in cases than is accounted for by a Poisson renewal model, and each pathogen exists on a spectrum of dictating the degree of overdisperseness (Lloyd-Smith et al., 2005): SARS, for example, is prone to many superspreading events (Shen et al., 2004); whereas pneumonic plague exhibits less variation in offspring cases (Lloyd-Smith et al., 2005).

To study the robustness of EpiCluster under more variable data generating processes, we generated data using the fast drop-off *R*_*t*_ profile and a negative binomial (NB) renewal model with inverse-dispersion parameter *κ >* 0: as *κ* → ∞, the NB model approaches the Poisson. So low values of *κ* correspond to more overdispersed data. Using the fast drop-off *R*_*t*_ profile, we generated case data under different values of *κ*, and, for each series, we fit the sliding window, EpiFilter and EpiCluster methods.

The results are shown in Fig. S5. When *κ* is large (i.e. the data are effectively generated from a Poisson distribution), the results match those observed in Fig. 3. As the data generating process exhibits more variation, all methods perform worse: generally failing to correctly identify the change in *R*_*t*_ and inferring a highly noisy *R*_*t*_ profile with many spurious fluctuations. However, the sliding window and EpiFilter methods generally produced more robust estimates in the presence of strong overdispersion.

### 3.4. EpiCluster estimates sharp changes in R_t_ for real COVID-19 incidence curves

Next, we performed retrospective inference of *R*_*t*_ for the early COVID-19 outbreaks in three selected regions: Victoria and Queensland, Australia, and Hong Kong (see 2.8), which were selected for the variety of transmission profiles they encompass. The *R*_*t*_ estimates for these regions are shown in Figure 5 again comparing the sliding window approach (panel B) with the EpiFilter approach (panel C) and EpiCluster (panel D).

**Figure 5:**
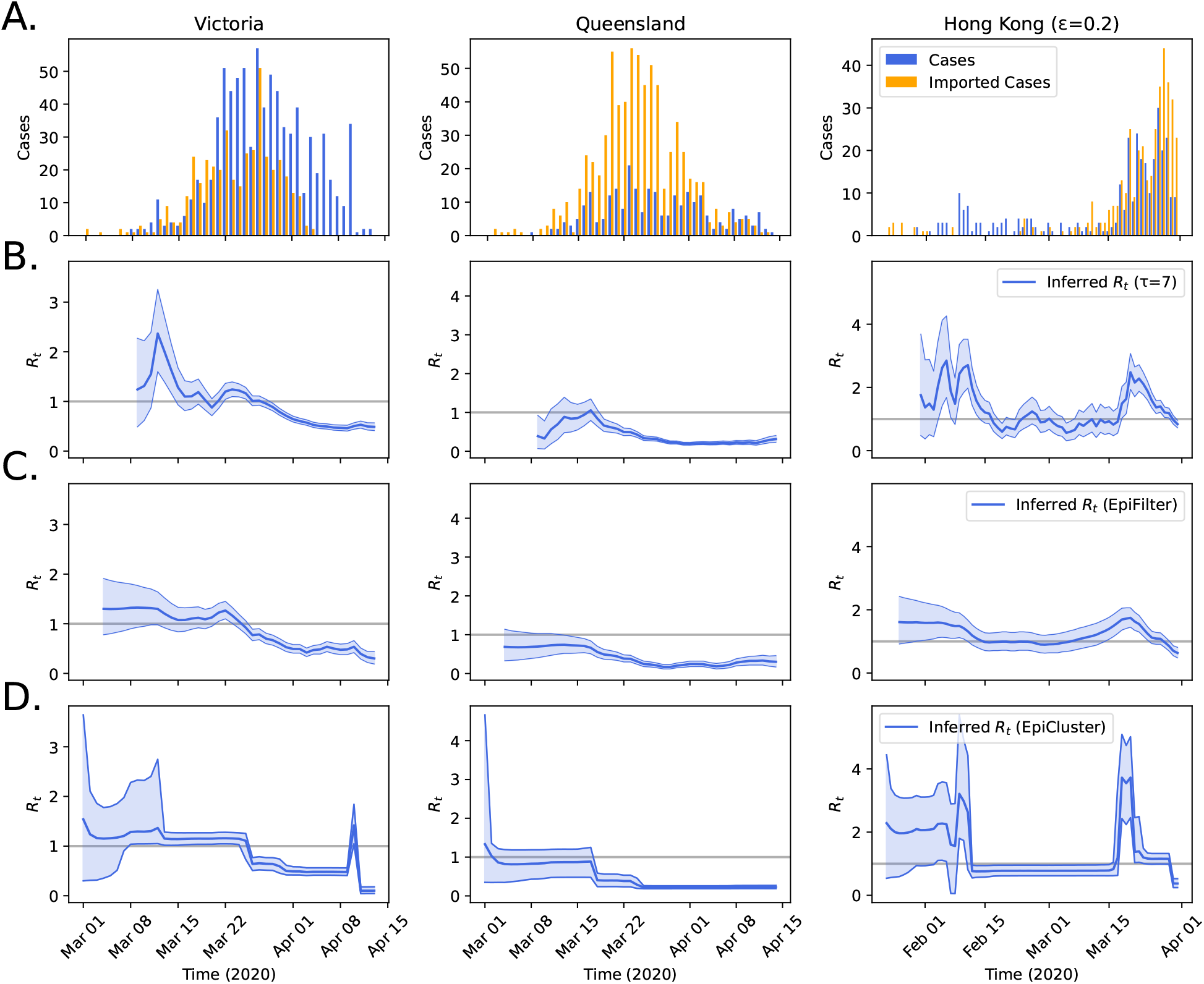
Learning R_*t*_ from early COVID-19 epidemic incidence curves in three locations. Data on local and imported cases from the early COVID-19 pandemic in three selected regions is shown in panel A. In panel B, we show the inferred R_*t*_ profile using a sliding window method (Thompson et al., 2019) for two different choices of the sliding window size (τ = 7 and 28 days). In panel C, we show the inferred R_*t*_ profile using the EpiFilter method (Parag, 2021). In panel D, we show the inference results when using EpiCluster to recover R_*t*_. In panels B, C and D, shaded regions indicate the central 90% of the posterior distribution of R_*t*_, while the central line indicates the posterior mean, and the background gray line indicates R_*t*_ = 1.

The first case of COVID-19 in Australia was reported in Victoria state on 25th January 2020 (Storen and Corrigan, 2020). Subsequently, Victoria quickly became a hub of transmission and declared a state of emergency on 16th March, including a ban on non-essential gatherings of over 500 people (Storen and Corrigan, 2020). On 18th March, more restrictions on movement followed with indoor public gatherings of more than 100 people banned and restrictions in aged care facilities introduced across Australia (Storen and Corrigan, 2020). On the 22nd March, the state Premier announced that Victoria would implement a shutdown of all non-essential activity across the state (Storen and Corrigan, 2020). The sliding window approach (Fig. 5B) and EpiFilter (Fig. 5C) both estimated declines in transmission starting around 22nd March; EpiCluster infers a sharper decline around 25th March. All methods inferred that transmission subsequently remained below the level for sustained transmission, apart from an uptick in transmission estimated from EpiCluster coinciding with a burst of cases around 10th April, which likely reflects a violation of the assumptions of the model.

The first case of COVID-19 in Queensland, Australia occurred on 29th January 2020 (Storen and Corrigan, 2020), and the first wave began in early March. All three estimation methods inferred that, since imported cases were the dominant cause of the wave, there was relatively low community transmission, and the bulk of local *R*_*t*_ estimates were below 1 (Fig. 5). All methods inferred a decline in transmission beginning around the 16th March—the date when Victoria declared a state of emergency—and EpiCluster estimated a rapid decline on 17th March. To combat the insurgence of imported cases, the Queensland Premier announced that the state would restrict access to the border on 24th March: this included termination of all rail services and border road closures (Storen and Corrigan, 2020), and EpiCluster inferred a small decline occurring on this date.

Hong Kong, like Singapore and Taiwan, was quick to act on learning of the outbreak of COVID-19 in Wuhan, China, and the government enacted intensive surveillance campaigns and declared a state of emergency on 25th January, 2020 (Cowling et al., 2020, Fig. 1). On the 7th February 2020, Hong Kong introduced prison sentences for anyone breaching quarantine rules (OT&P Healthcare, 2022). This date broadly coincides with the decline in *R*_*t*_ detected across all three methods, and the decline detected by EpiCluster is especially rapid.

Hong Kong’s second wave of COVID-19 began in March 2020 driven by imported cases from North America and Europe (Parag et al., 2021), and all three methods detect an increase in the local *R*_*t*_ shortly after 15th March. Policy responses to this wave by the Hong Kong government included a ban on foreign travellers (effective 25th March; OT&P Healthcare (2022)) and a ban on gatherings of more than four people (effective 27th March; OT&P Healthcare (2022)); a significant decrease in *R*_*t*_ is detected by all three methods around the times when these interventions were imposed. The EpiCluster results mirror the timing of this intervention most closely, suggesting that there was a short time lag between when the interventions were imposed and their effect.

To explore the sensitivity of our estimates for Hong Kong to the hyperparameters of the method, we performed a series of sensitivity analyses where these parameters were fixed at different values and inference was performed (Fig. S6). These experiments illustrate that, as either of the hyperparameters are increased, the *R*_*t*_ profile comprises a greater number of regimes, and there is greater uncertainty in the *R*_*t*_ estimates. The qualitative behaviour of the majority of estimates, however, remains the same, with a large decline in transmission around 7th February 2020 and a resurgence in mid March.

## 4. Discussion

The time-varying reproduction number, *R*_*t*_, is a threshold metric for facilitating decision making during epidemics. But, there is also value in using *R*_*t*_ estimates to retrospectively assess whether the imposition of interventions caused substantive and rapid reductions in transmission (e.g. Dehning et al., 2020; Flaxman et al., 2020). It is especially key to determine the timing of these reductions, since delays in imposition of interventions can substantially worsen outcomes, particularly during the growth of an epidemic (Pei et al., 2020). Here, we present a general Bayesian inference method using Pitman-Yor process priors which allows any feasible number of changepoints in transmission, and we provide a choice of hyperparameters (see §2.2.3), such that, *a priori, R*_*t*_ is assumed to remain relatively stable. Through simulated data examples, we show that the method is adept at estimating sharp changes in transmission: in both retrospective and real-time analyses. By fitting the model to real data from COVID-19 outbreaks, we infer discontinuous declines in transmission at times which broadly coincide with the imposition of interventions. The method allows effectively automated detection of changepoints in transmission and could be adapted to handle different types of models in epidemiology and, more generally, provides a framework for handling time-varying parameters.

The information available to estimate *R*_*t*_ changes throughout an epidemic: at the start, there is scant information, and estimates have high uncertainty; when an epidemic is brought under control, cases are initially higher, providing more information of changes in transmission; and resurgences qualitatively mirror the conditions at the start of an epidemic meaning *R*_*t*_ has greater uncertainty (Parag and Donnelly, 2022). Priors thus affect estimates differently at different stages during an epidemic and, by extension, variously for different types of epidemic. The fits of our model and the two comparator methods to COVID-19 case data demonstrate the strong information introduced by the priors. This makes sensitivity analyses particularly important, since no one prior choice satisfies all parties for all situations. We assume that *R*_*t*_ is piecewise-constant with transmission changing discontinuously with the number of pieces and location of breakpoints controlled through a Pitman-Yor process. If transmission changes more gradually, these assumptions are inappropriate, and a model which allows a more gradual change in *R*_*t*_ will perform better (e.g. EpiFilter; Parag, 2021). Similarly, if the model mischaracterises the data generating process, for example, by assuming that there are no substantial differences in transmissibility across individuals, estimates will also be poor (Fig. S5). Because of this, it is possible that the sharp changes in COVID-19 transmission identified by EpiCluster for the three locations considered (Fig. 5) reflected violations in the model’s assumptions, and future work is to adapt our framework to handle such processes. We did not consider reporting issues here, and these would likely also introduce biases (Gostic et al., 2020; Pitzer et al., 2021).

The Pitman-Yor process is an example from a broad class of models from Bayesian nonparametrics where the complexity of the models grows along with the volume and complexity of the data (Ghahramani, 2013). Gaussian processes belong also to this class (Rasmussen, 2003) and have found wide application across epidemiology, notably for producing geostatistical maps of disease prevalence for illnesses such as malaria (Bhatt et al., 2015). More data and data of greater variety and complexity are being routinely collected in epidemiological surveillance, and there is a host of Bayesian nonparametric models (e.g. those described in Griffiths and Ghahramani, 2011; Ghahramani, 2013), which are well-placed for their analysis.

Across epidemiology, discretely sampled data are used to infer continuous-time parameters, such as the time-varying reproduction number, *R*_*t*_ in outbreak analysis, the effective population size in phylogenetic Skyline models (Pybus et al., 2000) and the historical force of infection in catalytic models (Muench, 2013). In any of these cases, transmission can change abruptly, and a model such as ours could be used to identify periods of rapid change.

## Data Availability

Data for the Australian regions are available from the Australian national COVID-19 database (https://www.ncbi.nlm.nih.gov/pmc/articles/PMC7449695/); data for Hong Kong are available from the Hong Kong Department of Health COVID-19 database (https://data.gov.hk/en-data/dataset/hk-dh-chpsebcddr-novel-infectious-agent).

## 5. Acknowledgements

KVP acknowledges funding from the MRC Centre for Global Infectious Disease Analysis (reference MR/R015600/1), jointly funded by the UK Medical Research Council (MRC) and the UK Foreign, Commonwealth & Development Office (FCDO), under the MRC/FCDO Concordat agreement and is also part of the EDCTP2 programme supported by the European Union.

## S1. Supplementary materials

**Figure S1:**
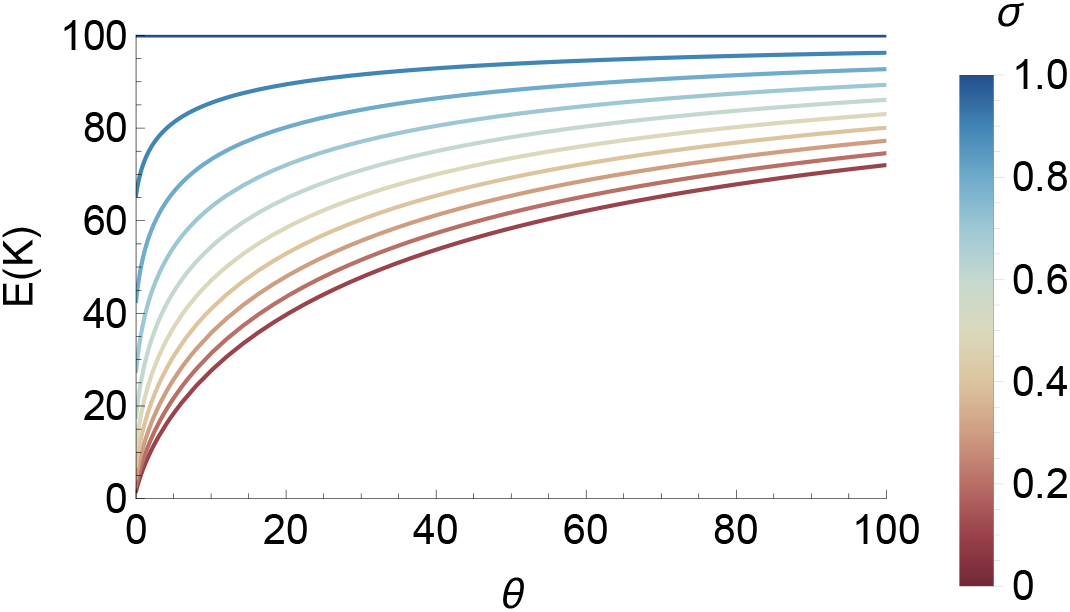
Expectation over the number of regimes (K) as a function of hyperparameters: θ and σ. This plot graphs eq. (5) and assumes that there are T = 100 datapoints.

**Figure S2:**
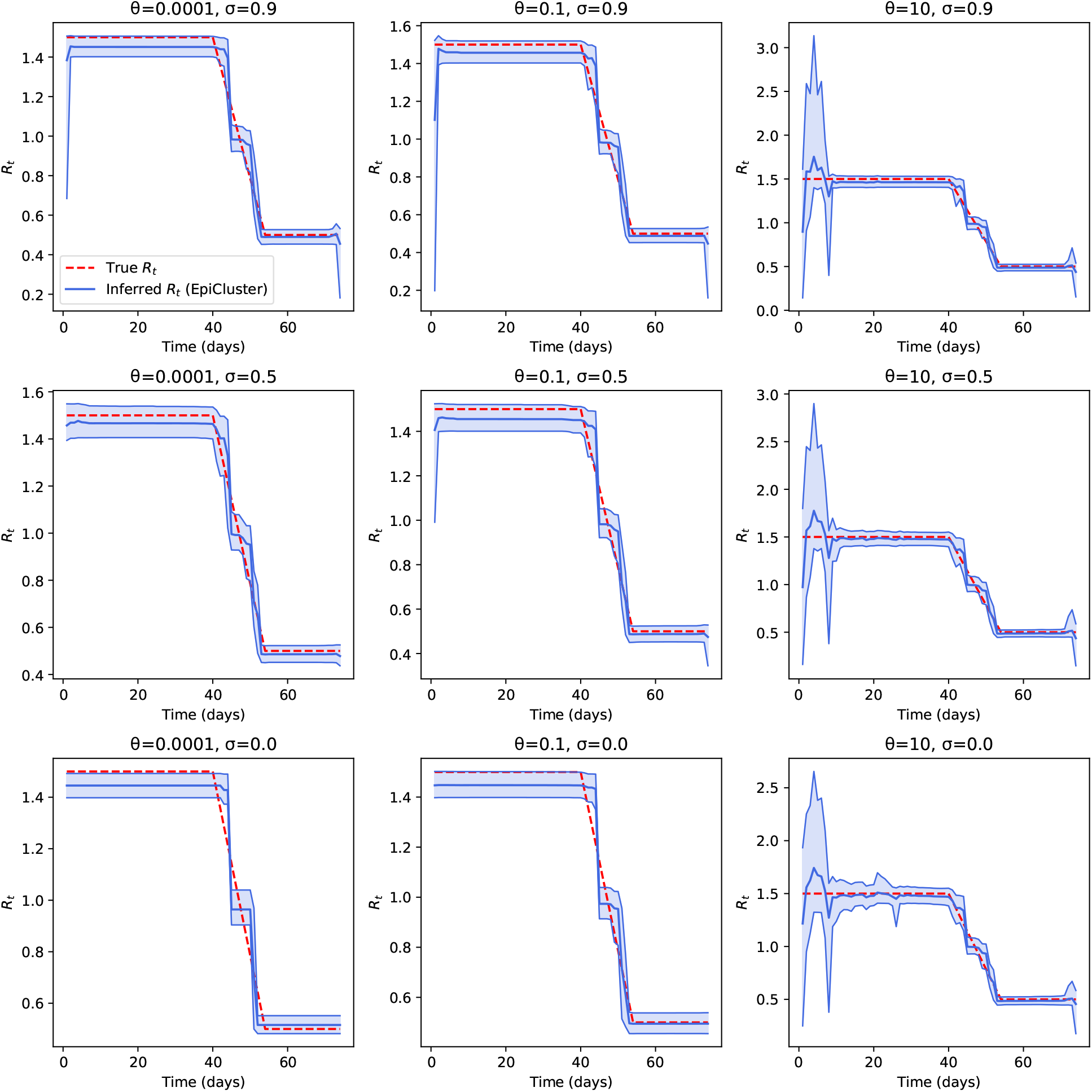
Effect of θ and σ on inference for R_*t*_. We used the same slow drop-off synthetic data from Figure 2, and performed inference for R_*t*_ using the indicated fixed values of θ and σ, the two hyperparameters of the Pitman-Yor process (see eq. (4)). In all panels, shaded regions indicate the central 90% of the posterior distribution of R_*t*_, while the central line indicates the posterior mean.

**Figure S3:**
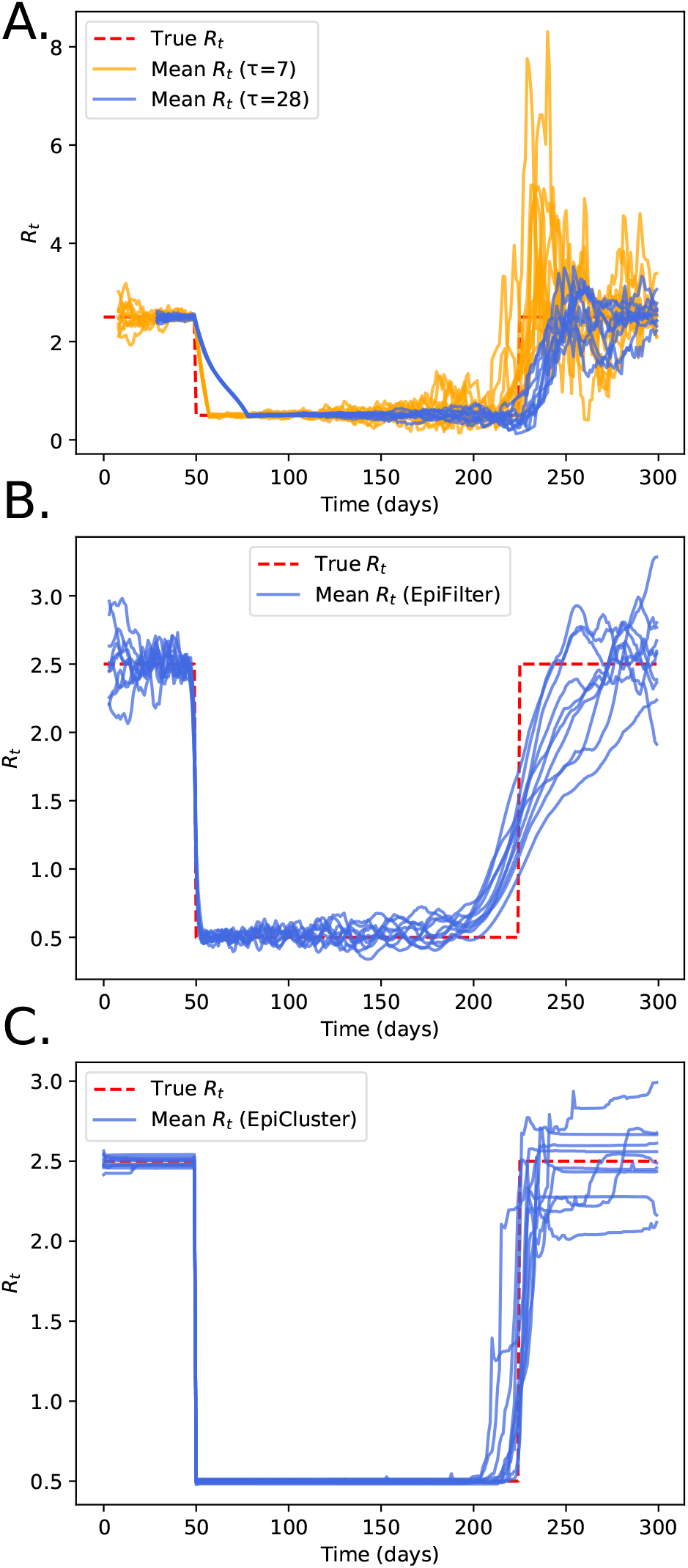
Posterior mean estimates of R_*t*_ in the fast resurgence synthetic example. Using the fast resurgence R_*t*_ profile (Fig. 2), we repeated the generation of synthetic data 10 times and performed inference for R_*t*_ for each synthetic dataset. Panel A shows the posterior means according to the sliding window method (Thompson et al., 2019) for two different choices of the sliding window size (τ = 7 and 28 days). In panel B, we show the inferred mean R_*t*_ profiles using the EpiFilter method (Parag, 2021). In panel C, we show the inferred means using EpiCluster.

**Figure S4:**
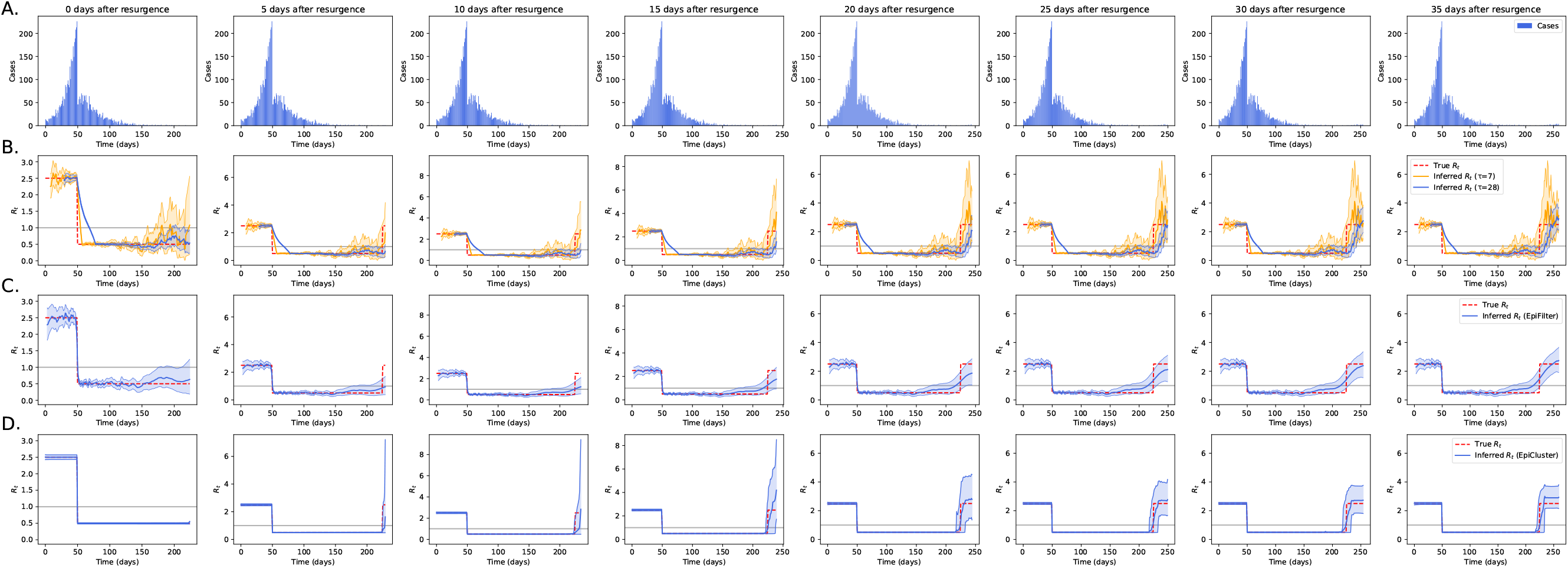
Real-time estimation of a resurgence in R_*t*_. We used the same fast resurgence synthetic data from Figure 2, and performed inference for R_*t*_ based only on the data indicated at the top of each column. In panel B, we show the inferred R_*t*_ profile using a sliding window method (Thompson et al., 2019) for two different choices of the sliding window size (τ = 7 and 28 days). In panel C, we show the inferred R_*t*_ profile using the EpiFilter method (Parag, 2021). In panel D, we show the inference results when using EpiCluster to recover R_*t*_. In panels B, C and D, shaded regions indicate the central 90% of the posterior distribution of R_*t*_, while the central line indicates the posterior mean, and the background gray line indicates R_*t*_ = 1.

**Figure S5:**
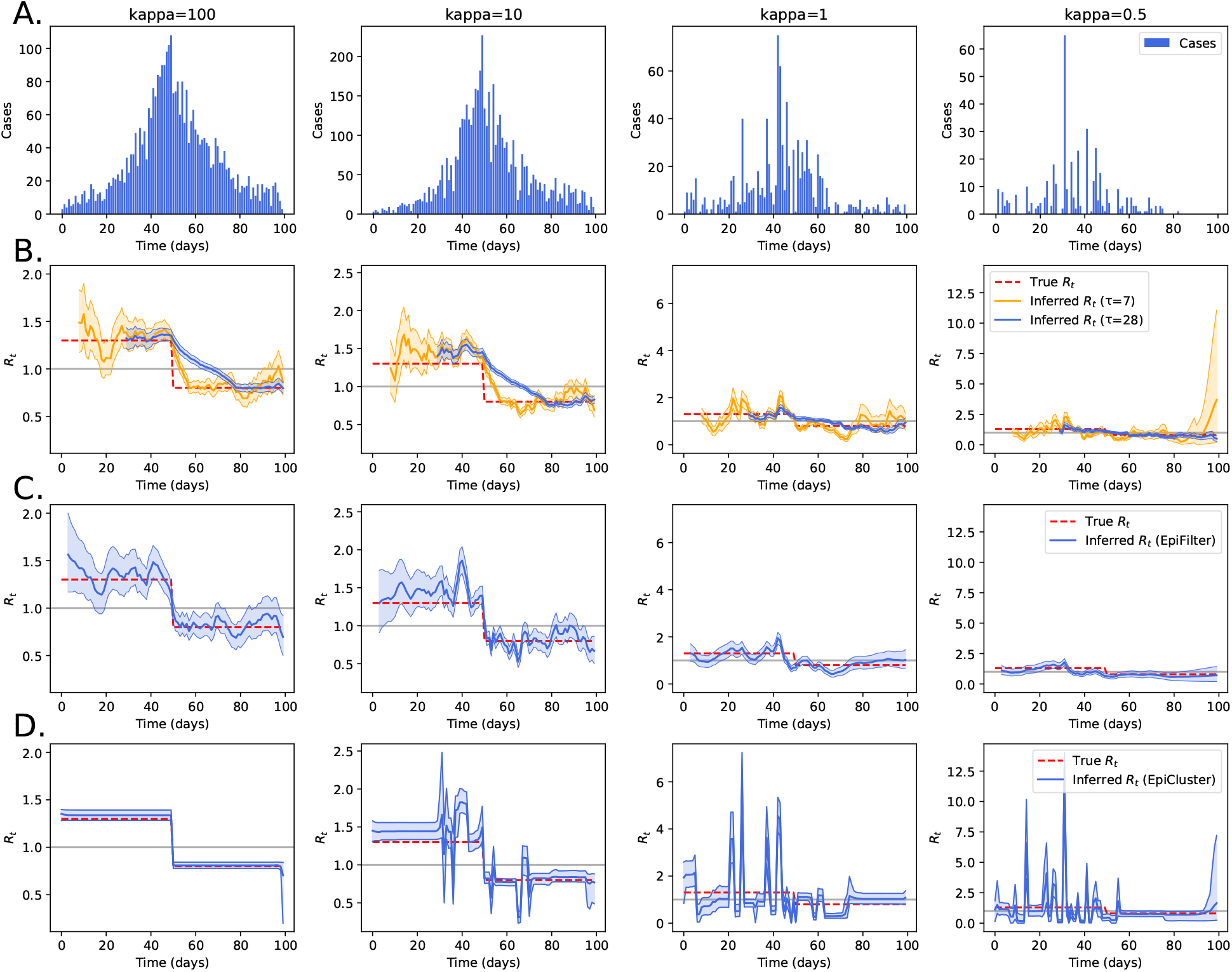
Effect of negative binomial noise on posterior estimates of R_*t*_. We used the same fast drop-off R_*t*_ profile from Figure 2 but generated data according to a negative binomial renewal process with the inverse overdispersion (kappa) indicated at the top of each column. In panel B, we show the inferred R_*t*_ profile using a sliding window method (Thompson et al., 2019) for two different choices of the sliding window size (τ = 7 and 28 days). In panel C, we show the inferred R_*t*_ profile using the EpiFilter method (Parag, 2021). In panel D, we show the inference results when using EpiCluster to recover R_*t*_. In panels B, C and D, shaded regions indicate the central 90% of the posterior distribution of R_*t*_, while the central line indicates the posterior mean, and the background gray line indicates R_*t*_ = 1.

**Figure S6:**
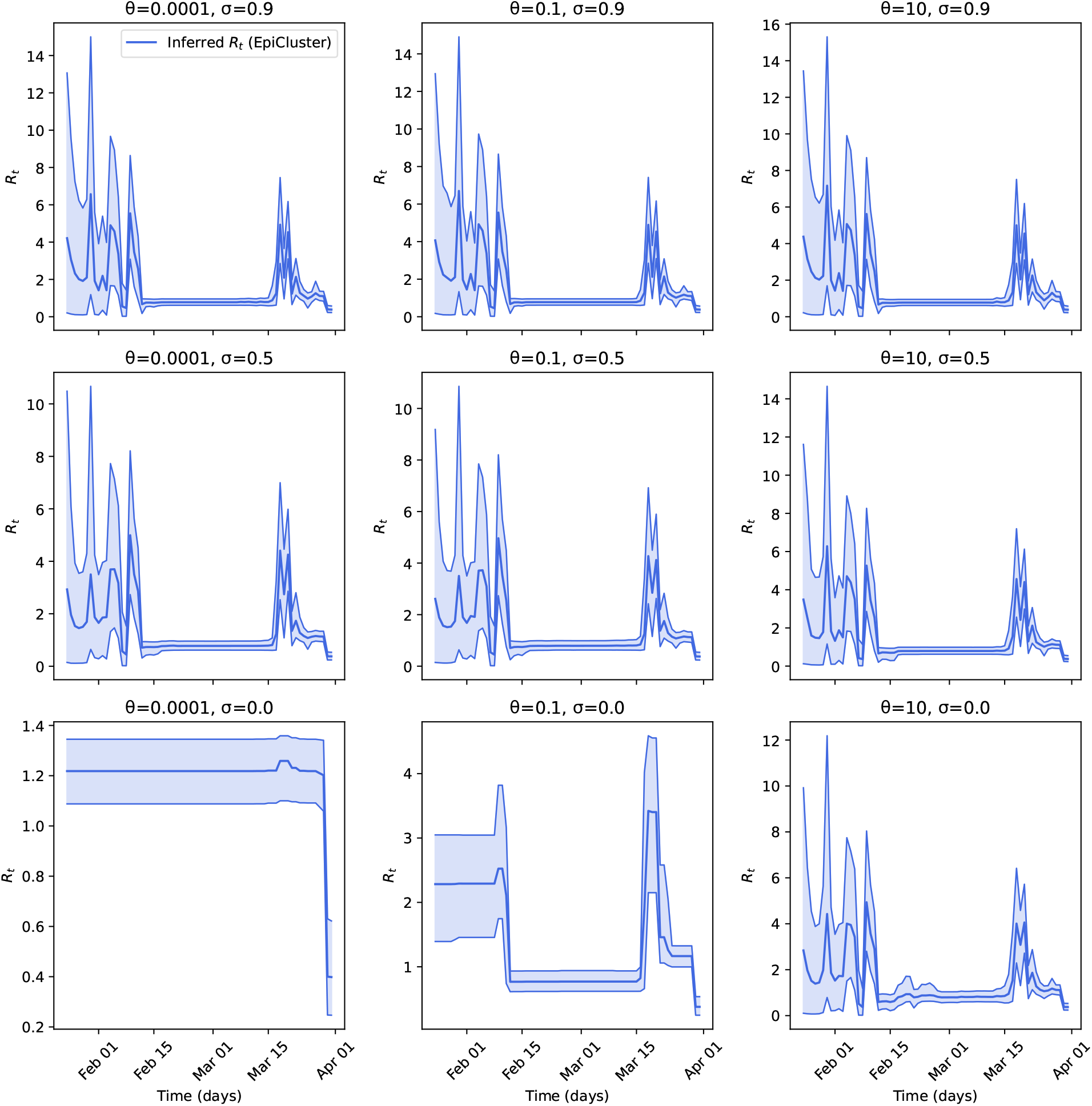
Effect of θ and σ on inference for R_*t*_ for the Hong Kong COVID-19 dataset. We used the Hong Kong data from Figure 4, and performed inference for R_*t*_ using the indicated fixed values of θ and σ, the two hyperparameters of the Pitman-Yor process (see eq. (4)). In all panels, shaded regions indicate the central 90% of the posterior distribution of R_*t*_, while the central line indicates the posterior mean.

## Footnotes

1 Also known as the effective reproduction number.

2 Technically, the renewal equation is formulated in terms of infections rather than cases, but, since we use the serial interval distribution in place of the generation time distribution, we keep with defining I_*t*_ as a case count.

3 Also known as the two-parameter Poisson-Dirichlet process.

4 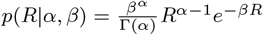.

## References

Abbott, S., Hellewell, J., Thompson, R.N., Sherratt, K., Gibbs, H.P., Bosse, N.I., Munday, J.D., Meakin, S., Doughty, E.L., Chun, J.Y., et al., 2020. Estimating the time-varying reproduction number of sars-cov-2 using national and subnational case counts. Wellcome Open Research 5, 112.

Bhatt, S., Weiss, D., Cameron, E., Bisanzio, D., Mappin, B., Dalrymple, U., Battle, K., Moyes, C., Henry, A., Eckhoff, P., et al., 2015. The effect of malaria control on plasmodium falciparum in africa between 2000 and 2015. Nature 526, 207–211.

Brauner, J.M., Mindermann, S., Sharma, M., Johnston, D., Salvatier, J., Gavenčiak, T., Stephenson, A.B., Leech, G., Altman, G., Mikulik, V., et al., 2021. Inferring the effectiveness of government interventions against covid-19. Science 371.

Cori, A., Ferguson, N.M., Fraser, C., Cauchemez, S., 2013. A new framework and software to estimate time-varying reproduction numbers during epidemics. American Journal of Epidemiology 178, 1505–1512.

Cowling, B.J., Ali, S.T., Ng, T.W., Tsang, T.K., Li, J.C., Fong, M.W., Liao, Q., Kwan, M.Y., Lee, S.L., Chiu, S.S., et al., 2020. Impact assessment of non-pharmaceutical interventions against coronavirus disease 2019 and influenza in hong kong: an observational study. The Lancet Public Health 5.

Creswell, R., Augustin, D., Bouros, I., Farm, H.J., Miao, S., Ahern, A., Robinson, M., Lemenuel-Diot, A., Gavaghan, D.J., Lambert, B., Thompson, R.N., 2022. Heterogeneity in the onwards transmission risk between local and imported cases affects practical estimates of the time-dependent reproduction number. Philosophical Transactions of the Royal Society A.

Dehning, J., Zierenberg, J., Spitzner, F.P., Wibral, M., Neto, J.P., Wilczek, M., Priesemann, V., 2020. Inferring change points in the spread of COVID-19 reveals the effectiveness of interventions. Science 369.

Flaxman, S., Mishra, S., Gandy, A., Unwin, H.J.T., Mellan, T.A., Coupland, H., Whittaker, C., Zhu, H., Berah, T., Eaton, J.W., Molod, M., Imperial College COVID-19 Response Team, Ghani, A.C., Donnelly, C., Riley, S., Vollmer, M.A.C., Ferguson, N.M., Okell, L.C., Bhatt, S., 2020. Estimating the effects of non-pharmaceutical interventions on COVID-19 in Europe. Nature 584, 257–261.

Fraser, C., 2007. Estimating individual and household reproduction numbers in an emerging epidemic. PLOS One 2.

Gelman, A., Rubin, D.B., 1992. Inference from iterative simulation using multiple sequences. Statistical Science, 457–472.

Ghahramani, Z., 2013. Bayesian non-parametrics and the probabilistic approach to modelling. Philosophical Transactions of the Royal Society A: Mathematical, Physical and Engineering Sciences 371.

Gostic, K.M., McGough, L., Baskerville, E.B., Abbott, S., Joshi, K., Tedijanto, C., Kahn, R., Niehus, R., Hay, J.A., De Salazar, P.M., et al., 2020. Practical considerations for measuring the effective reproductive number, Rt. PLOS Computational Biology 16.

Griffiths, T.L., Ghahramani, Z., 2011. The indian buffet process: An introduction and review. Journal of Machine Learning Research 12.

Hong Kong Department of Health, 2022. Latest local situation of COVID-19. https://data.gov.hk/en-data/dataset/hk-dh-chpsebcddr-novel-infectious-agent.

Lambert, B., 2018. A student’s guide to Bayesian statistics. Sage.

Li, Y., Campbell, H., Kulkarni, D., Harpur, A., Nundy, M., Wang, X., Nair, H., for COVID, U.N., et al., 2021. The temporal association of introducing and lifting non-pharmaceutical interventions with the time-varying reproduction number (r) of sars-cov-2: a modelling study across 131 countries. The Lancet Infectious Diseases 21, 193–202.

Lijoi, A., Prunster, I., 2010. Models beyond the Dirichlet process, in: Hjort, N.L., Holmes, C., Muller, P., Walker, S.G. (Eds.), Bayesian nonparametrics. Cambridge University Press.

Liu, Y., Gu, Z., Liu, J., 2021. Uncovering transmission patterns of COVID-19 outbreaks: A region-wide comprehensive retrospective study in Hong Kong. EClinicalMedicine 36.

Lloyd-Smith, J.O., Schreiber, S.J., Kopp, P.E., Getz, W.M., 2005. Superspreading and the effect of individual variation on disease emergence. Nature 438, 355–359.

Martínez, A.F., Mena, R.H., 2014. On a nonparametric change point detection model in Markovian regimes. Bayesian Analysis 9, 823–858.

Mendez-Brito, A., El Bcheraoui, C., Pozo-Martin, F., 2021. Systematic review of empirical studies comparing the effectiveness of non-pharmaceutical interventions against covid-19. Journal of Infection 83, 281–293.

Muench, H., 2013. Catalytic models in epidemiology, in: Catalytic Models in Epidemiology. Harvard University Press.

Nishiura, H., Chowell, G., 2009. The effective reproduction number as a prelude to statistical estimation of time-dependent epidemic trends, in: Mathematical and statistical estimation approaches in epidemiology. Springer, pp. 103–121.

Nishiura, H., Linton, N.M., Akhmetzhanov, A.R., 2020. Serial interval of novel coronavirus (COVID-19) infections. International Journal of Infectious Diseases 93, 284–286.

OT&P Healthcare, 2022. Covid-19 timeline of events. https://www.otandp.com/covid-19-timeline. Accessed: 22 June 2020.

Parag, K.V., 2021. Improved estimation of time-varying reproduction numbers at low case incidence and between epidemic waves. PLOS Computational Biology 17.

Parag, K.V., Cowling, B.J., Donnelly, C.A., 2021. Deciphering early-warning signals of sars-cov-2 elimination and resurgence from limited data at multiple scales. Journal of the Royal Society Interface 18.

Parag, K.V., Donnelly, C.A., 2020. Adaptive estimation for epidemic renewal and phylogenetic skyline models. Systematic Biology 69, 1163–1179.

Parag, K.V., Donnelly, C.A., 2022. Fundamental limits on inferring epidemic resurgence in real time using effective reproduction numbers. PLOS Computational Biology 18, e1010004.

Pei, S., Kandula, S., Shaman, J., 2020. Differential effects of intervention timing on covid-19 spread in the united states. Science Advances 6.

Pitman, J., 2002. Combinatorial stochastic processes. Lecture notes in mathematics 1875, 7–24.

Pitman, J., Yor, M., 1997. The two-parameter Poisson-Dirichlet distribution derived from a stable subordinator. The Annals of Probability, 855–900.

Pitzer, V.E., Chitwood, M., Havumaki, J., Menzies, N.A., Perniciaro, S., Warren, J.L., Weinberger, D.M., Cohen, T., 2021. The impact of changes in diagnostic testing practices on estimates of covid-19 transmission in the united states. American Journal of Epidemiology 190, 1908–1917.

Price, D.J., Shearer, F.M., Meehan, M.T., McBryde, E., Moss, R., Golding, N., Conway, E.J., Dawson, P., Cromer, D., Wood, J., Abbott, S., McVernon, J., McCaw, J.M., 2020. Early analysis of the Australian COVID-19 epidemic. eLife 9.

Pybus, O.G., Rambaut, A., Harvey, P.H., 2000. An integrated framework for the inference of viral population history from reconstructed genealogies. Genetics 155, 1429–1437.

Rasmussen, C.E., 2003. Gaussian processes in machine learning, in: Summer school on machine learning, Springer. pp. 63–71.

Roberts, M.G., Nishiura, H., 2011. Early estimation of the reproduction number in the presence of imported cases: pandemic influenza h1n1-2009 in new zealand. PLOS One 6.

Sharma, M., Mindermann, S., Brauner, J., Leech, G., Stephenson, A., Gavenčiak, T., Kulveit, J., Teh, Y.W., Chindelevitch, L., Gal, Y., 2020. How robust are the estimated effects of nonpharmaceutical interventions against covid-19? Advances in Neural Information Processing Systems 33, 12175–12186.

Shen, Z., Ning, F., Zhou, W., He, X., Lin, C., Chin, D.P., Zhu, Z., Schuchat, A., 2004. Superspreading sars events, beijing, 2003. Emerging infectious diseases 10, 256.

Soltesz, K., Gustafsson, F., Timpka, T., Jaldén, J., Jidling, C., Heimerson, A., Schön, T.B., Spreco, A., Ekberg, J., Dahlström, Ö., et al., 2020. On the sensitivity of non-pharmaceutical intervention models for sars-cov-2 spread estimation. medRxiv.

Storen, R., Corrigan, N., 2020. Covid-19: a chronology of state and territory government announcements (up until 30 june 2020). https://www.aph.gov.au/About_Parliament/Parliamentary_Departments/Parliamentary_Library/pubs/rp/rp2021/Chronologies/COVID-19StateTerritoryGovernmentAnnouncements#_Toc52275800. Accessed: 22 June 2020.

Svensson, Å., 2007. A note on generation times in epidemic models. Mathematical Biosciences 208, 300–311.

Teh, Y.W., 2010. Dirichlet process. Encyclopedia of Machine Learning 1063, 280–287.

Thompson, R., Stockwin, J., van Gaalen, R.D., Polonsky, J., Kamvar, Z., Demarsh, P., Dahlqwist, E., Li, S., Miguel, E., Jombart, T., Lessler, J., Cauchemez, S., Cori, A., 2019. Improved inference of time-varying reproduction numbers during infectious disease outbreaks. Epidemics 29.

Van Kerkhove, M.D., Bento, A.I., Mills, H.L., Ferguson, N.M., Donnelly, C.A., 2015. A review of epidemiological parameters from Ebola outbreaks to inform early public health decision-making. Scientific Data 2, 1–10.

Wallinga, J., Teunis, P., 2004. Different epidemic curves for severe acute respiratory syndrome reveal similar impacts of control measures. American Journal of Epidemiology 160, 509–516.

